# Genetic association analysis of 269 rare diseases reveals novel aetiologies

**DOI:** 10.1101/2022.06.10.22276270

**Authors:** Daniel Greene, Genomics England Research Consortium, Daniela Pirri, Karen Frudd, Ege Sackey, Mohammed Al-Owain, Arnaud P.J. Giese, Khushnooda Ramzan, Itaru Yamanaka, Nele Boeckx, Chantal Thys, Bruce D. Gelb, Paul Brennan, Verity Hartill, Julie Harvengt, Tomoki Kosho, Sahar Mansour, Mitsuo Masuno, Takako Ohata, Helen Stewart, Khalid Taibah, Claire L.S. Turner, Faiqa Imtiaz, Saima Riazuddin, Takayuki Morisaki, Pia Ostergaard, Bart Loeys, Hiroko Morisaki, Zubair M. Ahmed, Graeme M. Birdsey, Kathleen Freson, Andrew Mumford, Ernest Turro

## Abstract

The genetic aetiologies of more than half of rare diseases remain unknown^1^. Standardised genome sequencing (GS) and phenotyping of large patient cohorts provides an opportunity for discovering the unknown aetiologies^2^, but this depends on efficient and powerful analytical methods^3^. We have developed a portable computational and statistical framework for inferring genetic associations with rare diseases. At its core lies the ‘Rareservoir’, a compact database of rare variant genotypes and phenotypes. We built a Rareservoir of 77,539 genomes sequenced by the 100,000 Genomes Project (100KGP)^4^. We then applied the Bayesian association method, BeviMed^3^, across 269 rare diseases assigned to participants in the project, identifying 238 known^5^ and 21 novel associations. The novel results included three which we selected for validation. We provide compelling evidence that (1) loss-of-function variants in the ETS-family transcription factor encoding gene *ERG* lead to primary lymphoedema, (2) truncating variants in the last exon of TGFβ regulator *PMEPA1* result in Loeys-Dietz syndrome^6^, and (3) loss-of-function variants in *GPR156* give rise to recessive congenital hearing impairment. These novel findings confirm the power of our analytical approach for the aetiological discovery of rare diseases.

---

Collectively, rare diseases affect 1 in 20 people^7^, but fewer than half of the approximately 10,000 catalogued rare diseases have a resolved genetic aetiology^1^. Standardised GS of large, phenotypically diverse collections of rare disease patients enables aetiological discovery across a wide range of pathologies^2^. However, the scale and complexity of large GS datasets and the hierarchical nature of patient phenotype coding^8^ induces numerous bioinformatic and statistical challenges. Most importantly, the full genotype data from GS studies of tens of thousands of individuals are typically stored in unmodifiable variant call format (VCF) files many terabytes in size, leading to high storage and processing costs. Recently developed frameworks such as Hail^9^ and OpenCGA^10^ afford greater flexibility. However, they are designed to capture genotypes for variants across the full minor allele frequency (MAF) spectrum, from rare (MAF<0.1%) to common (MAF>5%) variants. To accommodate large numbers of genotypes, they depend on distributed storage systems and require numerous software packages, hindering deployment.

## The ‘Rareservoir’

We developed a compact and portable relational database schema, the ‘Rareservoir’, which enables efficient statistical analysis of rare variant genotypes and clinical phenotypes. A Rareservoir stores genotypes sparsely and for rare variants only—by default, those for which all population-specific MAFs are likely to be <0.1%. This greatly reduces the number of stored genotypes in a large study (**Extended Data Fig. 1**), allowing the data to reside in a relational database on centralised storage. Variants in this MAF stratum encompass the vast majority of those having a large effect on rare disease risk. The Rareservoir encodes variants as 64-bit integers (‘RSVR IDs’, **Fig. 1a**), which can encode 99.2% of variants encountered in practice without loss of information. RSVR IDs occupy a single column and increase numerically with respect to genomic position, allowing fast location-based queries within a simple database structure. To support the build process of a Rareservoir, we developed a complementary software package called ‘rsvr’ (**Fig. 1a, Extended Data Fig. 2**). The package includes tools to annotate variants with MAF information from control databases (e.g. gnomAD^11^), pathogenicity scores (e.g. CADD scores^12^) and predicted Sequence Ontology (SO)^13^ consequences with respect to a set of known transcripts. We use a 64-bit integer (‘CSQ ID’) to record the consequences for interacting variant/transcript pairs, where each bit encodes one of the possible consequences, ordered by severity. Encoding the consequences in this way is efficient and enables succinct queries that threshold or sort based on severity of impact. The Rareservoir also contains a table with genetically derived data for each sample (including ancestry, sex and membership of a maximal set of unrelated participants (MSUP)), and a table of ‘case sets’ storing the rare diseases assigned to each participant.

**Fig. 1.**
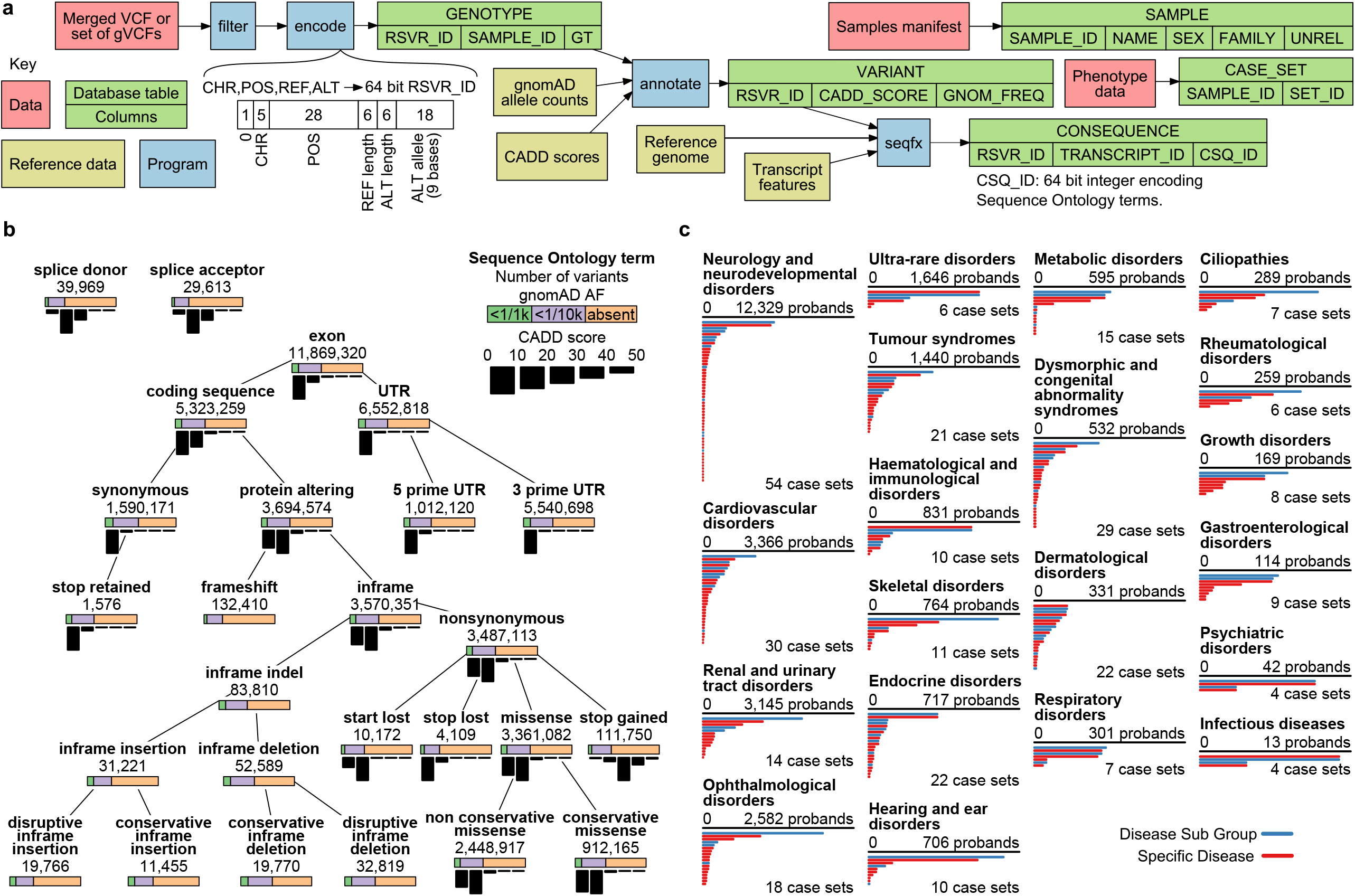
The Rareservoir in the 100KGP. **a**, Schematic of the database build procedure and contents. Variants are extracted from VCF files, filtered on internal cohort allele frequency, encoded as 64-bit RSVR IDs and loaded into a table containing the corresponding genotypes. The variants are annotated with scores reflecting their predicted deleteriousness (e.g. CADD score) and their gnomAD probabilistic minor allele frequency scores^2^. The consequences of each variant with respect to a reference set of transcripts are generated and loaded into a table. Sample information including pedigree membership and membership of an MSUP is loaded into a table. The case groupings for case/control association analyses are stored in a table. **b**, Schematic showing the variant data in the 100KGP Main Programme Rareservoir. The number of variant/transcript pairs, the distribution of CADD scores and a breakdown of gnomAD frequency classes is shown for each annotated SO term in the context of the structure of the ontology. **c**, Bars showing the size of each case set used for the genetic association analyses, grouped by Disease Group and coloured by type (Disease Sub Group or Specific Disease). Case sets smaller than 5 are shown as having size 4 to comply with 100KGP policy on limiting participant identifiability.

## BeviMed identifies 238 known and 21 novel genetic associations

We built a Rareservoir of 11.9 million rare exonic and splicing single nucleotide variants (SNVs) and short insertions or deletions (indels) in canonical transcripts of protein-coding genes in Ensembl v104^14^ from a merged VCF containing genotype calls for 77,539 participants in the Rare Diseases Main Programme of the 100KGP (Data Release Version 13) (**Fig. 1b**). During enrolment, expert clinicians assigned eligible cases to one or more of 220 Specific Diseases, arranged into 88 Disease Sub Groups, each of which belonged to one of 20 Disease Groups. We generated 269 case sets corresponding to the distinct Specific Diseases and Disease Sub Groups, ranging in size from 5,809 to one proband, and encoded them in the Rareservoir (**Fig. 1c, Extended Data Fig. 3**). We included these two levels of the phenotyping hierarchy to account for heterogeneity in presentation or diagnosis among cases sharing the same genetic aetiology, with the aim of boosting power to identify statistical genetic associations.

Using BeviMed, we obtained a posterior probability of association (PPA) between each of the 19,663 protein-coding genes and each of the 269 disease case sets. We selected probands in a given case set on the basis of pedigree information provided by the 100KGP and compared them to participants not in the case set who belonged to different pedigrees and to an MSUP, also provided by the 100KGP. To guard against false positive associations due to errors in the pedigree data and cryptic relatedness, we ensured the PPAs were robust to removal of cases sharing an excess of rare variants in the Rareservoir relative to ancestry-specific distributions. To account for correlation between case sets, we only recorded the association for each gene having the highest PPA within a given Disease Group. Conditional on a gene being causal for a particular disease, we recorded posterior probabilities over the mode of inheritance (MOI), the consequence class of variants that mediate disease risk (e.g. predicted loss-of-function variants or variants in the 5′ UTR) and the pathogenicity of each specific variant. Using a significance threshold of PPA>0.95, we identified 259 significant associations, 238 of which were documented by the PanelApp gene panel database^5^ with high, medium or low levels of prior supporting evidence in the literature (**Fig. 2, Supplementary Table 1**). These results give an upper bound on the false discovery rate (FDR) of 8%. In contrast, a recent analysis of 57,000 samples in the 100KGP reported 249 known and 579 novel associations^15^, giving an upper bound on the FDR of 70%, which suggests that our analytical approach has a greater specificity for a given sensitivity. Out of the 238 known associations that we identified, 40 (16.8%) were with Disease Sub Groups. This demonstrates that participants with different Specific Diseases belonging to the same Disease Sub Group sometimes share defects in the same gene, which confirms that treating Disease Sub Groups, not just Specific Diseases, as case sets, boosts statistical power.

**Fig. 2.**
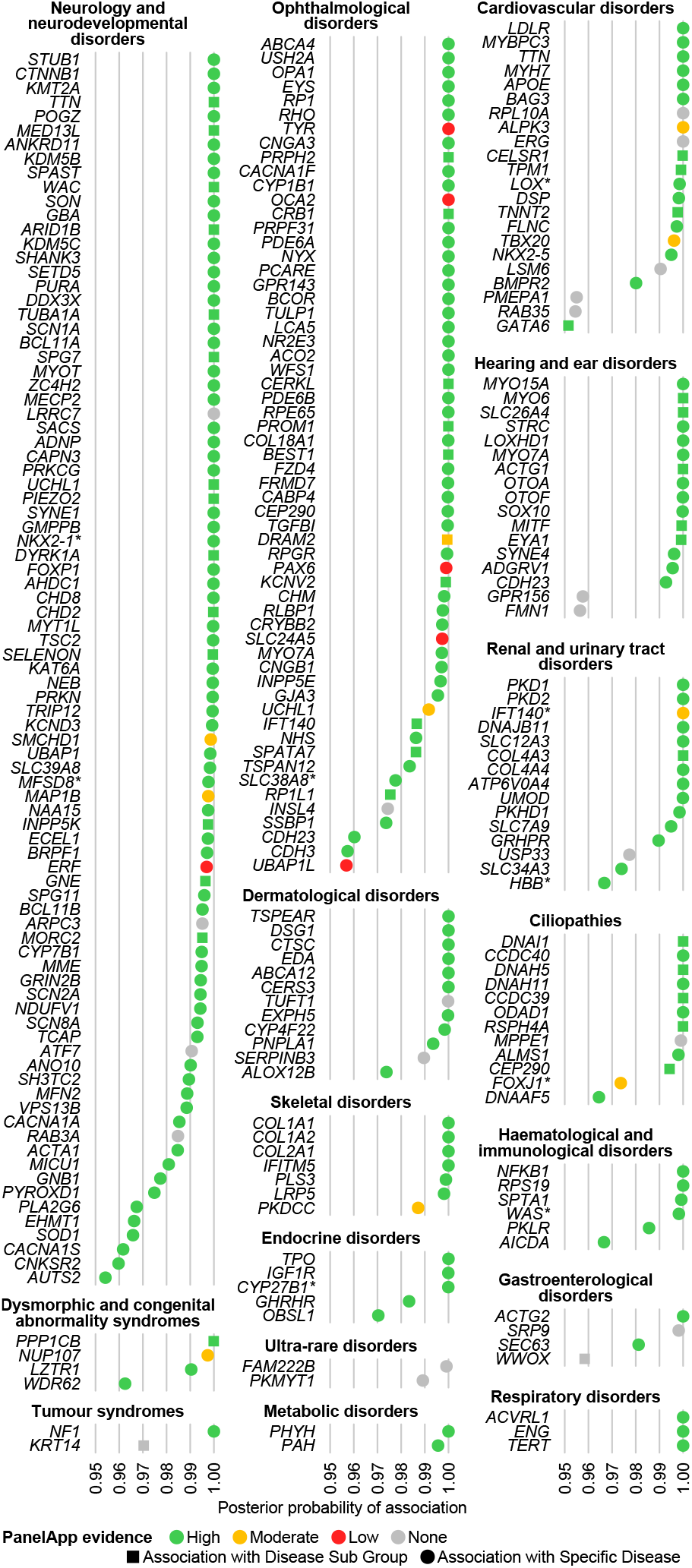
BeviMed genetic association results. BeviMed PPAs >0.95 arranged by Disease Group. Only the strongest association for each gene within a Disease Group is shown. Associations are coloured by their PanelApp evidence level (green, amber or red). Associations with substantial supporting evidence from the literature, but which were not automatically mapped to PanelApp, are marked with an asterisk (**Supplementary Table 1**). Novel associations are shown in grey. The shape of the points shows whether the association was with a Disease Sub Group (square), or Specific Disease (circle).

Out of the 238 associations identified as known according to PanelApp data, 234 (98.3%) had an inferred MOI that was consistent with the MOIs listed for the relevant gene in the matched panel (220 associations), in the notes for the matched panel (five associations) or in the MOIs listed for an alternative relevant panel (nine associations) in PanelApp (**Supplementary Table 1**). This provided independent evidence that the genetic associations we labelled as known (without reference to MOI information) are genuinely supported by evidence in the literature, further demonstrating the accuracy of BeviMed’s inference. Of the four known associations with an inferred MOI that was incongruous with PanelApp, two had supporting evidence for the inferred MOI in the literature that was absent from PanelApp: *EDA* with dominant ‘Ectodermal dysplasia without a known gene mutation’^16^ and *AICDA* with dominant ‘Primary immunodeficiency’^17^. The two associations with an MOI that was unsupported in the literature were between *UCHL1* and dominant ‘Inherited optic neuropathies’ and between *SLC39A8* and dominant ‘Intellectual disability’.

We found 21 novel genetic associations. To select a shortlist for further investigation, we developed a plausibility score (range 0–3) based on three sources of additional evidence (**Table 1**). Firstly, we considered evidence of purifying selection from gnomAD v2.1.1. Any dominant associations with high-impact variants in a gene having a probability of loss-of-function intolerance (pLI) >0.9 or with moderate-impact variants in a gene having a *Z*-score >2 were considered to be supported by population genetic metrics of purifying selection. To avoid disadvantaging recessive associations, which are unlikely to leave a detectable signature of purifying selection in gnomAD even if genuine, they were considered to be supported by default. Secondly, we considered co-segregation data: any association for which variants having a conditional posterior probability of pathogenicity >0.5 tracked with case status in at least three additional family members and for which no affected relatives lacked the pertinent variants were considered to be supported by co-segregation. Thirdly, we performed a comprehensive review of the literature for each gene and made a subjective assessment of whether an association was supported by biological function or previously known disease associations for related genes. In total, three genetic associations had a plausibility score of 3 and were therefore investigated further by considering additional experimental evidence or whether there was replication in other sequenced rare disease collections.

**Table 1.** Plausibility scoring of the 21 genetic associations identified by BeviMed. Each row corresponds to a genetic association between a gene and a case set in the 100KGP Main Programme without prior supporting evidence in PanelApp. Each column gives additional information for each association. Cells contributing to the final score are shown in bold. Associations with a score of three are shaded. The level of the case set in the disease label hierarchy (DSG: Disease Sub Group, SD: Specific Disease), the class of variants and the MOI identified as aetiological by BeviMed are shown (Dom: dominant; Rec: recessive). A recessive association contributes one point to the score. A pLI >0.9 contributes one point to the score providing the inferred class of aetiological variants is high-impact variants. A *Z*-score >2 contributes one point to the score providing the inferred class of aetiological variants is moderate-impact variants. Evidence of co-segregation in ≥3 relatives in the 100KGP data contributes one point to the score (including mosaicism supported by ≥2 reads containing the alternate allele). Prior evidence of a relevant biological function or disease association contributes one point to the score. The ‘Replication’ column specifies cohorts in which additional cases were confirmed.

| Gene | Case set | Level | Variant class | MOI | pLI | Z | Co-segregation evidence | Biological function and existing disease associations | Score | Replication |
| --- | --- | --- | --- | --- | --- | --- | --- | --- | --- | --- |
| *ERG* | Primary lymphoedema | SD | High | Dom | **0.96** | 2.53 | Co-segregation in 2 affected and 1 unaffected relatives (mosaicism in one affected parent). | ETS family transcription factor ERG is a critical regulator of endothelial lineage specification, vascular development, angiogenesis, and endothelial homeostasis.<sup>38,21</sup> | 3 | Internal (case enrolled for a different Specific Disease) |
| *GPR156* | Congenital hearing impairment | SD | High | Rec | 0 | 1.04 | Co-segregation in 2 affected and 4 unaffected relatives. | G protein-coupled receptor that regulates hair cell orientation in mechanosensory epithelia including in murine auditory epithelium<sup>32</sup>. | 3 | Maryland |
| *PMEPA1* | Familial Thoracic Aortic Aneurysm Disease | SD | High | Dom | **0.94** | 1.21 | Co-segregation in 3 affected relatives and distinctive phenotypic features. | Negative regulator of Transforming Growth Factor $\beta$ (TGFβ) signalling<sup>27</sup>. Aberrant TGFβ signalling is implicated in multiple Mendelian aortopathies<sup>28</sup>. | 3 | 100KGP pilot, Antwerp, Tokyo |
| *LRRC7* | Intellectual disability | SD | High | Dom | **1** | 3.6 |  | Brain-specific scaffold protein in post-synaptic densities<sup>35</sup>. *LRRC7*-inactivated mice have neuro-behavioural phenotype<sup>36</sup> | 2 |  |
| *USP33* | Extreme early-onset hypertension | SD | High | Dom | 0.86 | **2.1** | | Deubiquitinating enzyme implicated in multiple cellular processes, including regulation of expression of the β2-adrenergic receptor<sup>37</sup>, a critical regulator of circulatory function and blood pressure<sup>39</sup>. | 2 | |
| *ARPC3* | Charcot-Marie-Tooth disease | SD | Moderate | Dom | 0.22 | 0.39 |  | Component of the Arp2/3 complex that regulates polymerisation of F-actin, abundant in axonal neurofilaments. Multiple Mendelian axonal filamentopathies manifest as Charcot-Marie-Tooth disease<sup>40</sup>. *ArpC3*-inactivation in mice causes axon dysfunction<sup>41</sup>. | 1 |  |
| *ATF7* | Arthrogryposis | SD | High | Dom | **1** | 2.28 |  | Chromatin regulator implicated in innate immune response, adipocyte differentiation and telomerase regulation. | 1 |  |
| *FMN1* | Congenital hearing impairment | SD | High | Rec | 0 | -1.53 | Co-segregation in 2 unaffected relatives. | Formin family protein involved in linear actin and microtubule **polymerisation<sup>42</sup>. Pathogenic variants in the formin DIAPH1 cause hearing loss via cytoskeletal disruption in auditory stereocilia<sup>43</sup>.** | 1 |  |
| *RAB35* | Familial Hypercholesterolaemia | SD | High | Dom | **0.98** | 2.36 | Co-segregation in 1 affected relative. | Small GTP-binding proteins that are a regulator of endosomal transport and function. | 1 |  |
| *RAB3A* | Hereditary ataxia | SD | Moderate | Dom | 0.95 | **2.32** |  | Small GTP-binding proteins that regulate exocytosis and secretion. Although abundant in brain synaptic vesicles, rab3A-inactivated mice have no neuromuscular phenotype<sup>44</sup>. | 1 |  |
| *TUFT1* | Epidermolysis bullosa | SD | High | **Rec** | 0 | 0.9 | **Co-segregation in 1 affected and 4 unaffected relatives.** | Acidic protein that mediates dental enamel mineralisation. | 1 |  |
| *FAM222B* | Ultra-rare undescribed monogenic disorders | SD | Moderate | Dom | 0.29 | 0.42 |  | Uncharacterised nucleosomal protein. | 0 |  |
| *INSL4* | Rod Dysfunction Syndrome | SD | Moderate | Dom | 0 | -1.43 |  | Insulin-like growth factor implicated in trophoblast and bone development. | 0 |  |
| *KRT14* | Young onset tumour syndromes | DSG | Moderate | **Rec** | 0 | 0.84 | Co-segregation in 2 unaffected relatives. | Component of keratin intermediate filaments in epithelial cells. Pathogenic variants cause Epidermolysis bullosa simplex IA-D( AD/AR); Dermatopathia pigmentosa reticularis (AD); Naegeli-Franceschetti-Jadassohn syndrome (AD). | 0 |  |
| *LSM6* | Familial congenital heart disease | SD | High | Dom | 0.87 | 1.54 |  | Sm-like protein involved in spliceosome assembly and pre-mRNA splicing function. Putative regulator of mRNA degradation. | 0 |  |
| *MPPE1* | Primary ciliary dyskinesia | SD | High | **Rec** | 0 | 0.35 | Co-segregation in 2 unaffected relatives. | Metallophosphoesterase required for transport of GPI-anchor proteins from the endoplasmic reticulum to the Golgi. | 0 |  |
| *PKMYT1* | Single autosomal recessive mutation in rare disease | SD | Moderate | **Rec** | 0.22 | 0.07 | Co-segregation in 2 unaffected relatives. | Serine/threonine protein kinase that is a negative regulator of cell entry into mitosis. | 0 |  |
| *RPL10A* | Milroy disease | SD | High | Dom | 0.85 | 2.06 |  | Component of the large ribosomal subunit that mediates protein translation. | 0 |  |
| *SERPINB3* | Autosomal recessive congenital ichthyosis | SD | Moderate | **Rec** | 0 | -1.66 | Co-segregation in 2 unaffected relatives. | Cysteine endopeptidase inhibitor implicated in autocrine/paracrine signalling and cell protein metabolism | 0 |  |
| *SRP9* | Ductal plate malformation | SD | High | Dom | 0.42 | 1.13 |  | Component of the signal recognition particle that targets secretory proteins to the endoplasmic reticulum. | 0 |  |
| *WWOX* | Gastrointestinal disorders | DSG | Moderate | **Rec** | 0 | -4.44 | Co-segregation in 1 unaffected relative. | Short-chain dehydrogenase/reductase that acts as a tumour suppressor and apoptosis regulator. Pathogenic variants cause developmental and epileptic encephalopathy 28 and spinocerebellar ataxia 12. | 0 |  |

## Loss-of-function variants in *ERG* are responsible for primary lymphoedema

BeviMed identified a dominant genetic association between high-impact variants in *ERG* and the Specific Disease ‘Primary lymphoedema’. Primary lymphoedema is a group of genetic conditions caused by abnormal development of lymphatic vessels or failure of lymphatic function^18,19^. Three variants affecting different parts of the ERG protein were responsible for the high PPA, with locations ranging from codon 182 to 463 on the canonical Ensembl transcript ENST00000288319.12. One of the probands had two unaffected parents homozygous for the reference allele—one sequenced by the 100KGP and the other by Sanger sequencing—suggesting the truncating heterozygous variant had appeared *de novo*. A participant in a fourth family who had been enrolled to the 100KGP for an unrelated condition also carried a predicted loss-of-function variant in *ERG*. Upon manual chart review, this participant had features associated with this unrelated condition, but additional features consistent with primary lymphoedema, providing internal replication within the discovery cohort (**Fig. 3a**).

**Fig. 3.**
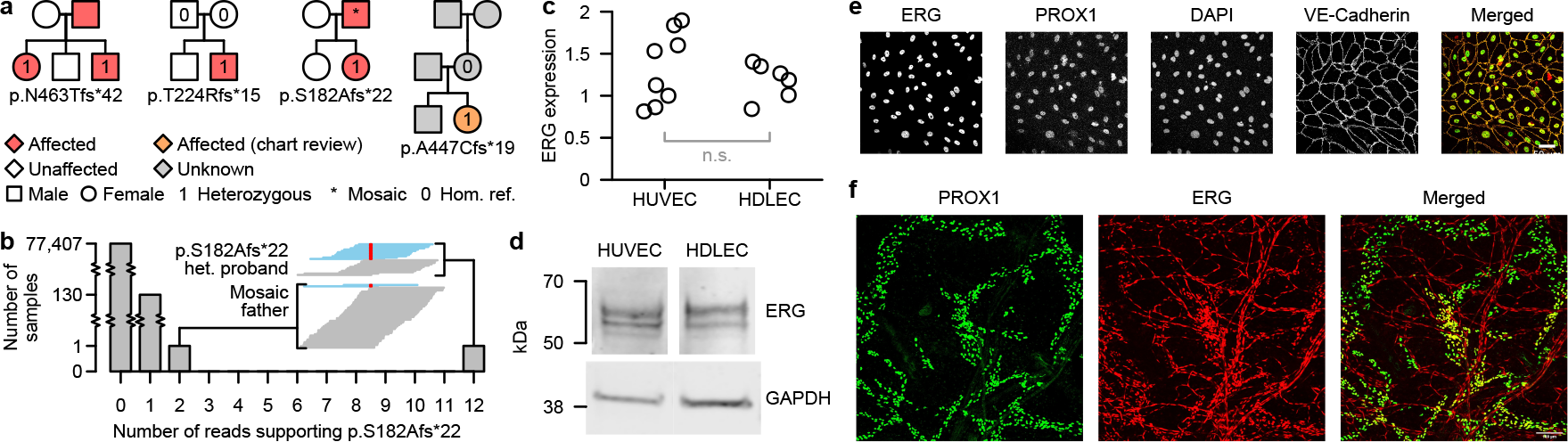
Loss-of-function variants in ERG are responsible for primary lymphoedema. **a**, Pedigrees for the four probands with loss-of-function variants in *ERG*: three were assigned the Specific Disease ‘Primary lymphoedema’ (left) and one was assigned to an unrelated disorder but later determined by chart review to also have lymphoedema (right-most pedigree). **b**, Truncated barchart showing the distribution of the number of reads supporting the p.S182Afs*22 alternate allele in the 100KGP. The embedded windows show the pileups of reads at this position in the two affected members of the family with the variant encoding p.S182Afs*22. The reads supporting the reference allele are in blue and those supporting the variant allele are in red. **c**, RT-PCR amplification of ERG mRNA in HDLEC relative to HUVEC. Data are normalised to GAPDH. Statistical significance was assessed using a Student’s *t*-test, n.s.: not significant. **d**, Representative immunoblot of HUVEC and HDLEC protein lysates identified several bands corresponding to ERG isoforms expressed at similar intensities in both cell types. **e**, Immunofluorescence microscopy of HDLEC shows ERG nuclear co-localisation with PROX1 and DAPI. HDLEC junctions are shown using an antibody to VE-cadherin. Scale bar, 50µm. **f**, *En face* immunofluorescence confocal microscopy of mouse ear skin. Vessels are stained with antibodies to the lymphatic marker PROX1 (green) and ERG (red). Scale bar, 100µm.

The affected father of the proband with the variant encoding p.S182Afs*22 was called homozygous for the reference allele, initially suggesting a lack of co-segregation of the variant with the disease in that pedigree. However, a review of the GS read alignments for the father revealed two independent reads overlapping that position which supported the alternative allele. To assess whether this could be the result of erroneous sequencing, we counted the number of such reads in the 77,539 genomes in the 100KGP and found that the proband and the father were the only two with more than one such read. This indicated that these reads in the father were unlikely to be erroneous but instead that he was mosaic. (**Fig. 3b**). This might explain why the father had a milder form of the disease, with an onset of lymphoedema over two decades later than his daughter.

ERG is a critical transcriptional regulator of blood vessel endothelial cell (EC) gene expression^20^ and is essential for normal vascular development^21^. However, little is known about the contribution of ERG to lymphatic development or how the loss-of-function variants identified in our analysis could result in primary lymphoedema. To compare the expression of ERG in lymphatic EC versus blood vessel EC, we carried out real-time quantitative PCR for *ERG* in RNA obtained from primary cultures of human umbilical vein EC (HUVEC) and human dermal lymphatic EC (HLDEC). No significant difference in *ERG* transcript levels were observed between HUVEC and HDLEC (**Fig. 3c**). Likewise, similar ERG expression was detected by immunoblotting of HUVEC and HLDEC protein lysates (**Fig. 3d**). Immunofluorescence microscopy was carried out to study the cellular localization of ERG in cultured HDLEC. These results confirmed nuclear co-localization of ERG with the lymphatic EC marker PROX1 (**Fig. 3e**). To understand whether ERG is present within the lymphatic vasculature *in vivo*, we analysed whole mount immunostaining of ERG and PROX1 in the ear skin of mice at three weeks after birth. At this time point, the lymphatic vessels have formed into a network of blind-ended lymphatic capillaries and valve-containing collecting vessels. We showed that ERG co-localizes with PROX1 in discrete lymphatic vascular structures that are distinct from the ERG signal present throughout the blood vessels in the mouse ear (**Fig. 3f**). Thus, we confirmed high levels of ERG expression in the lymphatic endothelium. In separate work, we have shown that deletion of the murine *ERG* homolog *Erg* in lymphatic endothelial cells results in defective lymphangiogenesis *in vivo*, confirming a role for *ERG* in lymphatic vessel remodelling and maintenance (Pirri et al., in preparation).

## Truncating variants in *PMEPA1* result in Loeys-Dietz syndrome

BeviMed identified a dominant genetic association between high-impact variants in *PMEPA1* and the Specific Disease ‘Familial Thoracic Aortic Aneurysm Disease’ (FTAAD). The variant with the highest conditional probability of pathogenicity was an insertion of one cytosine within a seven-cytosine stretch in the last exon of the canonical Ensembl transcript ENST00000341744.8. This variant, which is predicted to induce a p.S209Qfs*3 frameshift, was observed in three FTAAD pedigrees of European ancestry in the 100KGP discovery cohort. We replicated the association in three additional collections of cases. Firstly, the same variant was identified independently in eight affected members of three pedigrees of Japanese ancestry. Secondly, a single-cytosine deletion within the same poly-cytosine stretch as the previous variant, and encoding p.S209Afs*61, was found in an FTAAD case enrolled in a separate collection of 2,793 participants in the 100KGP Pilot Programme. Lastly, we identified a family in Belgium wherein the affected members carried a five base-pair deletion in the same stretch of poly-cytosines as above, inducing a frameshift two residues upstream of the other two variants (p.P207Qfs*3).

All pedigrees exhibited dominant inheritance of aortic aneurysm disease with incomplete penetrance and Marfanoid skeletal features with complete penetrance, which co-segregated with the respective variants in genotyped participants (**Fig. 4a**). To assess whether *PMEPA1* families affected by FTAAD form a phenotypically distinct subgroup, we analysed the HPO terms assigned to the 593 FTAAD families in both programmes of the 100KGP. Using a permutation-based method^22,23^ based on Resnik’s semantic similarity measure^24^, we found that the four 100KGP *PMEPA1* families were significantly more similar to each other than to other FTAAD families chosen at random (*p*=5.7×10^−3^). To characterise the *PMEPA1* phenotype in greater detail, we compared the prevalence of each of the HPO terms in the minimal set of terms present in at least three of the four families with the prevalence in the other FTAAD families. We identified four HPO terms related to the musculoskeletal system that were significantly enriched (**Fig. 4b**), echoing the phenotypic characteristics of the syndromic aortopathy Loeys-Dietz syndrome^25^. The cases in the Japanese and Belgian families also exhibited a constellation of skeletal phenotypes, including pectus deformity, scoliosis and arachnodactyly, consistent with observations in the UK families.

**Fig. 4.**
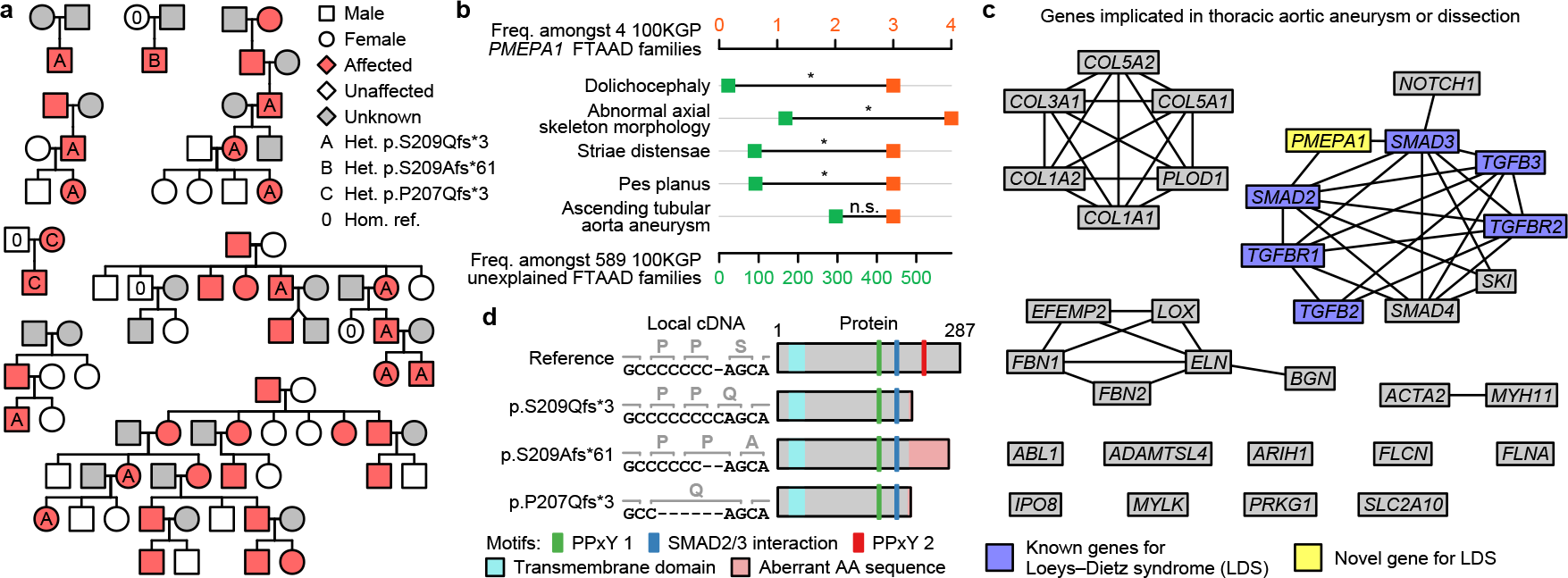
Truncating variants in *PMEPA1* result in Loeys-Dietz syndrome. **a**, Pedigrees for the three probands in the 100KGP (discovery cohort) heterozygous for the frameshift insertion encoding p.S209Qfs*3 and probands from replication cohorts, including: one from the 100KGP pilot programme heterozygous for the frameshift deletion encoding p.S209Afs*61, three of Japanese ancestry heterozygous for p.S209Qfs*3 and one Belgian pedigree heterozygous for a frameshift deletion encoding p.P207Qfs*3. **b**, HPO terms present in at least three of the four *PMEPA1* FTAAD families, excluding redundant terms within each level of frequency, alongside their frequency in four *PMEPA1* FTAAD families and the other 589 unexplained FTAAD families. Terms are ordered by *p*-value obtained by a Fisher’s exact test of association between the term’s presence in a FTAAD family and whether the family is one of the four *PMEPA1* families. Terms were declared significant (indicated by an asterisk), or not significant (n.s.) by comparing their Fisher test *p*-values and rank to a null distribution of equivalent pairs obtained by permutation (10,000 replicates). For each rank, the *p*-value of the term on the 5^th^ percentile was used as a threshold for declaring an association significant, provided all terms at higher ranks were also significant. **c**, Graph showing *PMEPA1* and genes with high evidence (green) of association with FTAAD in PanelApp. Edges connect genes where the string-db v11.5^26^ confidence score for physical interactions between corresponding proteins was >0.6. Genes known to be associated with Loeys-Dietz syndrome are highlighted in blue. *PMEPA1* is highlighted yellow. **d**, Schematic showing the effects of each variant at the cDNA and amino acid level, and on the protein product.

To understand the molecular mechanisms underlying this novel defect, we examined the protein-protein interactions^26^ among the complete set of high-confidence genes in the ‘Thoracic aortic aneurysm or dissection’ PanelApp panel and *PMEPA1*. PMEPA1 is a negative regulator of Transforming Growth Factor β (TGFβ) signalling^27^, which has previously been implicated in multiple aortopathies, including Loeys-Dietz syndrome^28^. The genes implicated in Loeys-Dietz syndrome form part of a tightly interacting subgroup of proteins in the TGFβ pathway, and PMEPA1 interacts with two of these: SMAD2 and SMAD3 (**Fig. 4c**). As the two candidate variants occur in the last exon of the transcript, they are likely to evade nonsense-mediated decay^29^. Their truncating effects are predicted to remove a PPxY interaction motif, while leaving the SMAD interaction motif intact (**Fig. 4d**), possibly affecting binding between PMEPA1 and SMAD2/3, and altering TGFβ signalling through a gain-of-function mechanism.

## Loss-of-function variants in *GPR156* give rise to recessive congenital hearing loss

BeviMed identified a recessive genetic association between high-impact variants in *GPR156* and the Specific Disease ‘Congenital hearing impairment’. Two high-impact variants in *GPR156* were responsible for the strong evidence of association: a one base pair deletion encoding p.S207Vfs*113 and a one base pair insertion encoding p.P718Lfs*86 with respect to the canonical Ensembl transcript ENST00000464295.6. In one family, each of the two parents heterozygous for p.S207Vfs*113 transmitted the frameshift allele to each of their two affected children, who were therefore homozygous for that allele. In a second family, the mother was heterozygous for the same p.S207Vfs*113 allele, which she transmitted to her two affected daughters. The father, however, was heterozygous for a different frameshift allele, encoding p.P718Lfs*86. He transmitted that allele to the same two affected daughters, who were thus compound heterozygous for truncating alleles. Using GeneMatcher^30^, we identified a third pedigree with biallelic truncating variants in *GPR156*. This pedigree concerned a consanguineous family with four siblings affected by hearing impairment, all of whom were homozygous for a variant encoding p.S642Afs*162 (**Fig. 5a**). The eight affected individuals in these three families all had congenital non-syndromic bilateral sensorineural hearing loss (see **Extended Data Fig. 4** for illustrative audiograms).

**Fig. 5.**
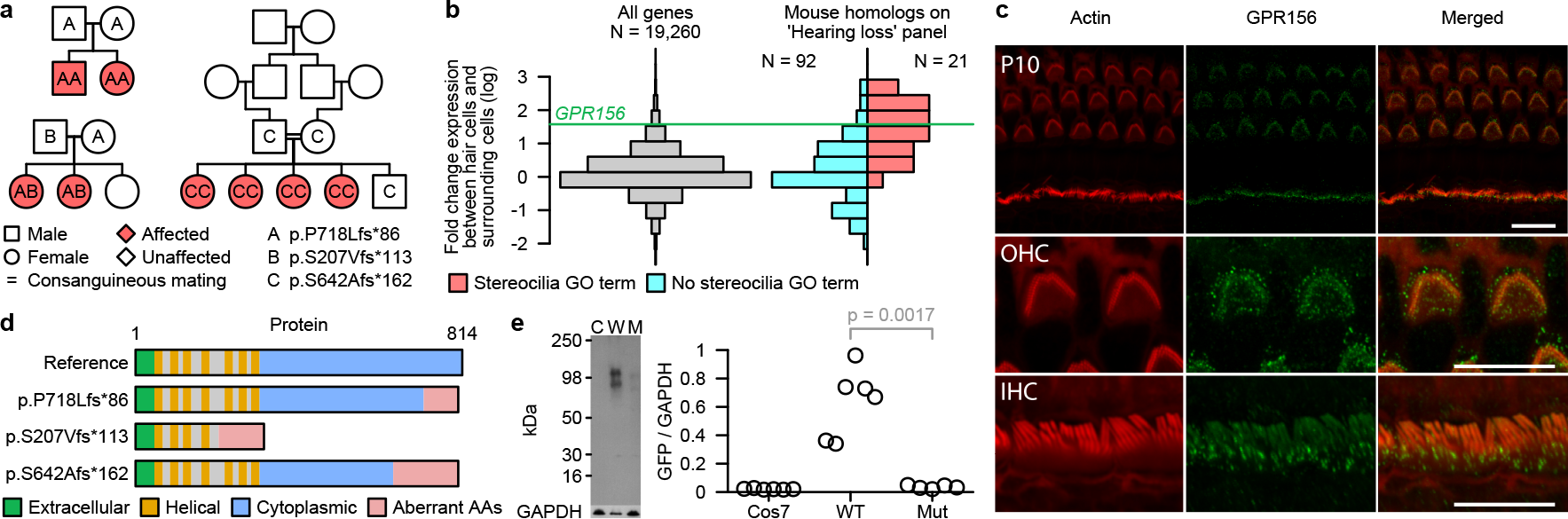
Loss-of-function variants in GPR156 give rise to recessive congenital hearing loss. **a**, Schematic of the three pedigrees with cases homozygous or compound heterozygous for loss-of-function variants in *GPR156*. Blank symbols indicate individuals with an unknown genotype. **b**, Histograms of expression log fold changes for different sets of genes in mouse hair cells compared to surrounding cells: all genes (left) and genes homologous to the human counterparts in the ‘Hearing loss’ PanelApp panel with and without a stereocilia-related GO term (i.e. a term whose name contained ‘stereocilia’ or ‘stereocilium’, or the descendant of such a term) (right). The log fold change for *Gpr156* is shown as a horizontal line. **c**, Maximum intensity projections of confocal Z-stacks in the organ of Corti and vestibular system of a P20 wild type mouse immunostained with GPR156 antibody (green) and counterstained with phalloidin (red). Top row: overview of the organ of Conti and vestibular system. Middle and bottom rows: magnified images of outer hair cells (OHC) and inner hell cells (IHC), respectively. No stereociliary bundle staining was observed. The punctate staining observed in the organ of Corti was absent or significantly decreased in the utricle of the vestibular system. Scale bars: 10 μm. **d**, Schematic showing the effects of each variant at the cDNA and amino acid level, and on the protein product. **e**, Representative western blot of GFP-GPR156 in untransfected Cos7 cells (C), Cos7 cells transfected with the wild type construct (W) and Cos7 cells transfected with the construct containing the mutant p.S642Afs*162 allele (M) (left). The ratios of GFP/GAPDH intensities in the three different conditions across technical replicates. The ratios corresponding to cells that had been transfected with the mutant construct were significantly lower than those corresponding to cells that had been transfected with the wild type construct (*p*=0.0017, *t*-test), but not significantly different from those corresponding to untransfected cells (*p*>0.05, *t*-test).

*GPR156* encodes a class C G protein-coupled receptor (GPCR) with sequence homology to the GABAB receptors^31^. Although previously designated as an orphan GPCR, *GPR156* has recently been identified as a critical regulator of stereocilia orientation on hair cells of the auditory epithelium and other mechanosensory tissues^32^. Its expression is highly restricted to hair cells in the inner ear^33^. Disruption of stereocilia is a common pathogenic mechanism underlying many human Mendelian hearing loss disorders^34^ and the over-expression of *GPR156* in hair cells relative to surrounding cells was commensurate with the over-expression of the 21 genes currently implicated in hearing impairment having a Gene Ontology (GO) term relating to stereocilia (**Fig. 5b**). By immunostaining of the Corti and vestibular system from P10 wild type mice, we found that GPR156 strongly co-localises with actin at the apical surface of the outer and inner hair cells of the organ of Corti (**Fig. 5c**).

Although one of the variants interrupted the sequence of seven transmembrane receptors of GPR156, two of the variants were located in the region encoding the GPCR’s cytoplasmic tail, within the gene’s last exon (**Fig. 5d**). To determine the effect of the truncating variants on GPR156 protein, we transfected Cos7 cells, which do not express human *GPR156* endogenously, with GFP-coupled constructs of wild type *GPR156* sequence and mutant *GPR156* sequence containing the p.S642Afs*162 allele. While cells transfected with wild type sequence expressed GFP-GPR156 protein robustly, cells transfected with the mutant construct did not express the protein appreciably, suggesting that the mutant protein is degraded and pointing to a loss-of-function mechanism (**Fig. 5e**). These data suggest that the biallelic chain truncating variants in *GPR156* cause a congenital hearing loss by preventing expression of GPR156 protein and thereby disrupting stereocilia formation in the auditory epithelium.

## Discussion

The standardisation of GS within a healthcare system, together with powerful frameworks for genetic and phenotypic data processing and statistical analysis, promises to advance the resolution of the remaining unknown aetiologies of rare diseases. We have developed a lightweight and easily deployable relational database, the Rareservoir, for genetic association analysis of rare diseases using approaches such as BeviMed. Of the 21 novel associations, we shortlisted, replicated and validated three. These three novel aetiologies, which we identified in a single unified analysis of one large cohort, involve genes that had not previously been implicated in any of these human diseases. The remaining 18 novel associations include further tantalising hypotheses. For example, *LRRC7*, which we identified to be associated with intellectual disability, encodes a brain-specific protein in post-synaptic densities^35^, and Lrrc7-deficient mice exhibit a neuro-behavioural phenotype^36^. *USP33*, which we found to be associated with early-onset hypertension, encodes a deubiquitinating enzyme implicated in regulating expression of the β2-adrenergic receptor regulation^37^. These and other candidates will require replication and validation before they can be considered causative genes. In addition, the exploration of rare variation in other regions of the genome using similar analytical approaches are a promising focus of future research.

## METHODS

### Ethics

The 100,000 Genomes project was approved by East of England–Cambridge Central REC REF 20/EE/0035. Only participants who consented for their data to be used for research were included in the analyses. The study at the University of Maryland was approved by the institutional review board (RAC#2100001) and written informed consent was obtained from the participating individuals. The study of the Japanese ancestry pedigrees bearing *PMEPA1* truncating alleles was approved by the Institutional Review Board of the National Cerebral and Cardiovascular Centre (M14-020) and Sakakibara Heart Institute (16-035), and written informed consent was obtained from the participating individuals.

### Building a Rareservoir

A Rareservoir is a relational database built through a series of steps from a set of input data and parameters (**Fig. 1a, Extended Data Fig. 2**). The ‘bcftools’ program^45^ extracts (‘bcftools view’) and normalises (‘bcftools norm’) variants from either a set of single sample gVCFs or from a merged VCF. In all steps of the procedure, variants are encoded as RSVR IDs using the ‘rsvr enc’ tool (see **Encoding RSVR IDs**). Merged VCFs typically contain cohort-wide variant quality information in the FILTER column, which can be used to select variants for processing. However, this is not readily obtained from single gVCFs. To address this, we developed the ‘rsvr depth’ tool, which computes variant quality pass rates at all positions in the genome based on a random subsample of gVCFs. If the input is a merged VCF, an internal (i.e. within-VCF) allele frequency threshold is applied with bcftools to filter out internally common variants. If the input is a set of single-sample gVCFs, internally common variants are filtered out in two steps, for computational efficiency. Firstly, a set of variants that are statistically almost certain to be common based on a random sample of gVCFs is identified—by default, the variants for which a one-sided binomial test under the null hypothesis that the MAF=0.01 is rejected at a significance level of 10^−6^ (done using the ‘rsvr tabulate’ tool). Secondly, all gVCFs are read sequentially, filtering out the variants identified in the previous step (using the ‘rsvr mix’ tool) and those for which the pass rates identified with ‘rsvr depth’ do not meet the threshold. Retained genotypes are then loaded into a temporary genotype table in the database in order to apply the final internal allele frequency filter by executing an SQL ‘DELETE’ statement. These variants are then annotated with gnomAD ‘probabilistic minor allele frequency’ (PMAF) scores^2^ using the ‘rsvr pmaf’ tool. The PMAF score is calculated with respect to a given allele frequency threshold *t*, by evaluating a binomial test (at a significance threshold of 0.05) on the observed frequency of the variant under the null hypothesis that the variant has an allele frequency of *t*. If, in any gnomAD population, the null is rejected for *t*=0.001 and the allele count is at least 2, the score is set to 0. If the null is rejected for *t*=0.0001, the score is set to 1. If the null is not rejected, the score is set to 2. Finally, if the variant is absent from gnomAD, the score is set to 3. For the non-pseudo autosomal dominant regions of chromosome X, only allele counts for males are used in calculations. Variants are then additionally annotated with their CADD phred scores using the ‘rsvr ann’ program and loaded into the VARIANT table. At this point, variants in the VARIANT and GENOTYPE table which have a PMAF score of 0 may be deleted because they are unlikely to be involved in rare diseases. We then annotate the retained variants with predicted transcript consequences for a given set of transcripts specified in a Gene Transfer Format (GTF) file. The ‘rsvr seqfx’ program determines a set of SO terms for each interacting transcript–variant pair and encodes them as a CSQ ID, which is added to the CONSEQUENCE table. The contents of the GTF file are also imported into the database to create tables of transcript features (FEATURE), transcripts (TX) and genes (GENE). Optionally, VARIANT, GENOTYPE and CONSEQUENCE may be filtered for RSVR IDs that have CSQ IDs meeting particular criteria, for instance, in order to retain only variants with protein-coding consequences. The SAMPLE table of metadata and genetic statistics for each sample represented in the input VCF(s) must then be added to the database, including mandatory columns containing the ID, sex, family, and an indicator of inclusion in the maximal unrelated set of samples in the database. The VARIANT, GENOTYPE and CONSEQUENCE tables are indexed by RSVR ID, in order to support fast lookups by genomic location. The SAMPLE table and GENOTYPE table are indexed by sample ID allowing fast lookups by sample. The CONSEQUENCE, TX and GENE tables are indexed by transcript and gene ID, allowing fast lookups of variants based on gene/transcript specific consequences. If sample phenotypes have been encoded using phenotypic terms (e.g. ICD10 codes or HPO terms), terms from the relevant coding systems can be added to a generic PHENOTYPE table mapping code IDs to descriptions, and codes assigned to samples can be added to the SAMPLE_PHENOTYPE table. Disease labels may be added to the CASE_SET table. The majority of the compute time required for building the database is taken by reading the genotype data from the input VCF files, which may be executed in parallel over separate regions against a merged VCF or over single gVCF files. The rsvr tool, implemented in C++, executes rapidly, with ‘rsvr seqfx’ capable of assigning CSQ IDs for all Ensembl v104 canonical transcripts to all variants (over 685M) in gnomAD v3.0 in under 40 minutes on a single core. Specific details on implementation of the workflow, code for encoding data as SQL statements compatible with Rareservoir and the mapping between bits in the 64 bit CSQ ID and each SO term assigned by ‘rsvr seqfx’ can be found in the rsvr software package (see **Code availability**).

### Encoding RSVR IDs

SNVs and indels may be encoded as 64-bit integers called RSVR IDs. In order to compute an RSVR ID for a given variant, the following expression is evaluated:

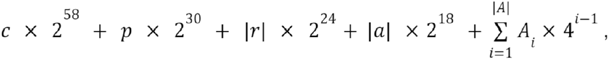

where *c* is the chromosome number (using 23, 24 and 25 respectively to represent X, Y and MT), *p* is the position, and |*r*| and |*a*| are the lengths of the reference and alternate alleles, respectively. *A* is a sequence identical to the alternate allele, *a*, when its length is less than 10, and otherwise equal to the first five followed by the last four elements of *a*. In the summation, nucleotides are assigned values: A= 0, C = 1, G = 2 and T = 3. The expression evaluates to integers that can be represented using 63 bits, setting the most significant bit to 0 when encoding as 64-bit integers. The chromosome, position, reference and alternate allele lengths and alternate allele bases are thereby encoded respectively by the subsequent 5, 28, 6, 6 and 18 bits (with two bits per base for the alternate allele). This procedure and its inverse are implemented in the ‘rsvr enc’ and ‘rsvr dec’ programs respectively. The proportion of variants in gnomAD 3.0 weighted by allele count that can be encoded losslessly is 99.2%, while 99.7% of all variants in gnomAD 3.0 can be represented by a distinct RSVR ID. Structural variants that can be represented by a position and length may also be encoded using distinct 64-bit RSVR IDs alongside SNVs and indels by setting the most significant bit to 1, and subsequently encoding the type of structural variant using 2 bits (Deletion 0, Duplication 1, Inversion 2, Insertion 3), the chromosome using 5 bits (as done for SNVs and indels), and the start and length consecutively using 28 bits.

### Genetic association analysis of 100KGP data

We constructed a Rareservoir in the Genomics England Research Environment containing the PASSing^46^ variants in the merged VCF file of 77,539 consented participants in the 100KGP rare diseases programme. Variants were filtered according to the default criteria described above using CADD v1.5 and gnomAD v3.0. Additionally, we imposed a minimum genotype quality of 35 and at least one predicted consequence on a canonical transcript in Ensembl v104. For each of the 269 case sets (**Extended Data Fig. 3**), we applied the BeviMed association test to rare variants extracted from the Rareservoir database in each of the 19,663 canonical transcripts belonging to a gene with a ‘protein_coding’ biotype. When we applied BeviMed, we averaged over models including the following classes of variant:

- 5’ UTR variants: those with a 5_prime_UTR_variant consequence (prior probability of association 0.0001),
- High impact variants: those with any consequences amongst start_lost, stop_lost, frameshift_variant, stop_gained, splice_donor_variant or splice_acceptor_variant (prior 0.00495),
- Moderate or high impact variants: those with any high impact consequence or missense_variant or inframe_deletion consequences (prior 0.00495).

Thus the prior probability of association was set to 0.01. To select the samples and set the case status used for each application of BeviMed to a disease–gene pair, one affected person per pedigree was selected, whilst the controls not from selected pedigrees were chosen from the MSUP. For the purposes of the association analysis, participants were labelled ‘explained’ by a given gene if they had variants in that gene classified as ‘pathogenic_variant’ or ‘likely_pathogenic_variant’ in the ‘gmc_exit_questionnaire’ table in the Research Environment. Case status was then set to one for affected individuals who were either not explained or explained by the target gene and zero for everyone else. Analyses yielding a BeviMed PPA >0.95 were re-run including all samples (i.e. with relatives). Associations for which the analysis with all samples caused the PPA to fall below 0.9 were filtered out, as the drop indicated a lack of co-segregation. To guard against false positives due to incorrect pedigree data, population structure or cryptic relatedness, we applied the following algorithm. Firstly, we obtained the distribution of the number of rare variants in the Rareservoir shared by pairs of individuals with a given ancestry in the 100KGP. The top percentile in each of these distributions was used to indicate potential relatedness between two cases in a given population. Analyses yielding a BeviMed PPA >0.95 were re-run after removing cases so as to ensure that any remaining set of cases carrying the same rare variant in the gene were not potentially related according to the criterion above. Associations for which this analysis caused the PPA to fall below 0.25 were filtered out.

### PanelApp annotation

Significant associations were coloured according to PanelApp^5^ (**Fig. 2**) evidence levels for panel–gene relations (green for high evidence, amber for moderate evidence, and red for low evidence) for panels of type ‘Rare Disease 100K’, which are organised hierarchically by Disease Sub Group and Disease Group, or of type ‘GMS Rare Disease’. Given an association between a gene and a case set (corresponding either to a Specific Disease or a Disease Sub Group), we searched for panels which contained the gene and had the same name as the case set (ignoring case). If such a match was not found, we searched for panels which contained the gene and which belonged to a Disease Sub Group with the same name as the Disease Sub Group of the case set. When this matching rule generated multiple matches, we selected the panel(s) with the highest evidence. If multiple panels still remained, we selected the panel with the smallest number of genes. Associations for which no matching in PanelApp could be found were inspected manually to assess whether PanelApp contained a suitable panel that could not be matched by the algorithm (marked with an asterisk in **Fig. 2**).

### Shortlisting novel genetic associations for validation

Several sources of independent evidence were used to shortlist significant associations for validation. For each source, a score of one was awarded if the evidence was supportive, and zero otherwise. Scores were then added over the different sources and used to rank the associations. Associations for which at least three sources of evidence were supportive were taken forward for further investigation. The sources of evidence and qualifying criteria for being considered supportive are listed below. Note that here we refer to variants which had a probability of pathogenicity >0.5 conditional on association as ‘probably pathogenic’.

- *Counting co-segregating pedigree members*. The pedigrees harbouring probably pathogenic alleles (1 for dominant associations and 2 for recessive) were checked for co-segregation between genotype and affection status. This evidence counted as supportive for associations for which all such pedigrees demonstrated co-segregation, and there were at least three additional relatives who had not been included in the association analysis but for whom there was co-segregation. Note that BAM files for the affected members of pedigrees who were called homozygous reference for probably pathogenic variants were checked for evidence of mosaicism to guard against the possibility that they were falsely portraying a lack of co-segregation.
- *pLI and Z-scores*. pLI and *Z*-scores for depletion of missense variants were obtained from the gnomAD v2.2.1 browser^11^. pLI >0.9 for associations in which high impact variants were most strongly associated were counted as supportive, whilst *Z*-scores greater than 2 for associations in which moderate impact variants were most strongly associated were counted as supportive.
- *Recessive association*. Population genetic metrics of purifying selection (pLI scores and *Z*-scores) are sensitive to depletion of high-impact variants and missense variants, respectively. They are therefore useful measures to corroborate dominant associations. However, these metrics have low sensitivity to identify the signatures of selection against recessive diseases because isolated pathogenic variants in heterozygous form do not lead to a reduction in reproductive fitness. To avoid disadvantaging recessive associations identified by BeviMed, they were assigned a contribution of one point to the score.
- *Literature review*. A comprehensive literature review, assessing the gene’s role (if any) in biological processes relevant to the disease, other diseases, and a survey of model organisms was undertaken, and determined to be either supportive or not.

### *ERG*: Primary endothelial cell culture

Single donor primary human dermal lymphatic endothelial cells (HDLEC) (Promocell, Heidelberg) were cultured in Endothelial Cell Growth Medium MV2 (Promocell). Pooled donor human umbilical vein endothelial cells (HUVEC) (Lonza, Slough) were grown in Endothelial Cell Growth Media-2 (EGM-2) (Lonza). HUVEC and HDLEC were grown on 1% (v/v) gelatin and used between passages 3-5.

### *ERG*: Real-time polymerase chain reaction

HUVEC and HDLEC were grown to confluency in a pre-gelatinised 6-well dish. Total RNA was isolated using the RNeasy Mini Kit (Qiagen) and 1 µg of total RNA was transcribed into cDNA using Superscript III Reverse Transcriptase (Thermo Fisher Scientific). Quantitative real-time PCR was performed using PerfCTa SYBR Green FastMix (Quanta Biosciences) on a Bio-Rad CFX96 System. Gene expression values of ERG in HUVEC and HDLEC were normalised to GAPDH expression and compared using the ΔΔCt method. The following oligonucleotides were used: ERG, 5’-GGAGTGGGCGGTGAAAGA-3’ and 5’-AAGGATGTCGGCGTTGTAGC-3’; GAPDH, 5’-CAAGGTCATCCATGACAACTTTG-3’ and 5’-GGGCCATCCACAGTCTTCTG-3’.

### *ERG*: Immunoblotting analysis

Immunoblotting was performed according to standard conditions. Proteins were labelled with the following primary antibodies: rabbit anti-human ERG antibody (1:1000; ab133264, Abcam) and mouse anti-human GAPDH (1:10000; MAB374, Millipore). Primary antibodies were detected using fluorescently labelled secondary antibodies: goat anti-rabbit IgG DyLight 680 and goat anti-mouse IgG Dylight 800 (Thermo Scientific). Detection of fluorescence intensity was performed using an Odyssey CLx imaging system (Li-COR Biosciences, Lincoln) and Odyssey ver.4 software.

### *ERG*: Immunofluorescence analysis of endothelial cells and mouse tissues

Confluent cultures of HUVEC and HDLEC were fixed with 4% (w/v) paraformaldehyde for 15 minutes and permeabilized with 0.5% (v/v) Triton-X100, before incubation with 3% BSA (w/v) in PBS containing the following primary antibodies: goat anti-human PROX1 antibody (1:100; AF2727, R&D Systems), rabbit anti-human ERG antibody (1:100; ab92513, Abcam), mouse anti-human VE-cadherin (1:100; 555661, BD Biosciences). Secondary antibody incubation was carried out in 3% BSA (w/v) in PBS, using the following antibodies: donkey anti-goat IgG Alexa Fluor-488 (1:1000; A-11055), donkey anti-rabbit IgG Alexa Fluor-555 (1:1000; A-31572), donkey anti-mouse Alexa Fluor-594 (1:1000; A-21203). All secondary antibodies from Thermo Fisher Scientific. Nuclei were visualized using DAPI (4′,6-diamidino-2-phenylindole) (Sigma-Aldrich). Confocal microscopy was carried out on a Carl Zeiss LSM780 confocal laser scanning microscope with Zen 3.2 software. All animal experiments were conducted with ethical approval from Imperial College London under UK Home Office Project Licence number PEDBB1586 in compliance with the UK Animals (Scientific Procedures) Act of 1986. Ear tissue was collected from euthanised 3-week old male and female C57BL/6J mice and fixed in 4% (w/v) paraformaldehyde at room temperature for 2h. Tissue was then washed with PBS followed by a blocking and permeabilization step using 3% (w/v) milk in PBST (containing 0.3% (v/v) Triton X-100 in PBS) for 1h at room temperature. The following primary antibodies were used for immunofluorescence staining: goat anti-human PROX1 antibody (1:100; AF2727, R&D Systems) and rabbit anti-human ERG antibody (1:100; ab92513, Abcam). Primary antibodies were incubated at 4 °C overnight in 3% (w/v) milk in PBST. The following day, tissues were washed three times with PBST over the course of 2h at room temperature. Tissues were incubated with secondary antibodies at room temperature for 2h in 3% milk (w/v) in PBST. Primary antibodies were detected using fluorescently labelled secondary antibodies: donkey anti-goat IgG Alexa Fluor-488 (1:400; A-11055, Thermo Fisher Scientific) and donkey anti-rabbit IgG Alexa Fluor-555; A-31572, Thermo Fisher Scientific). Stained samples were mounted onto glass slides using Fluoromount G (Thermo Fisher Scientific). Images were acquired using Zeiss LSM-780 confocal laser scanning microscope with Zen 3.2 software. All confocal images represent maximum intensity projection of Z-stacks of single tiles.

### *GPR156*: Western blot

We subcloned *GPR156* from human brain cDNA, into EGFP-C1 vector. The mutant *GPR156* construct was generated by mutagenesis using the QuickChange kit (Stratagene, La Jolla, CA) and a wild type GFP-GPR156 as a template. For expression analysis, the WT and mutant GPR156 experiments were transfected in COS7 cells grown in DMEM (Gibco, Gaithersburg, MD, USA) with 10% fetal bovine serum. Transfections were performed with Lipofectamine 2000 reagent (Life Technologies). Cells were harvested 24hr after transfection, lysed in buffer containing 1% CHAPS, 100mM NaCl, and 25mM HEPES, pH 7.4 and clarified by centrifugation at 14,000 rpm. Lysates (20μg) were run on a 15% SDS-PAGE gel. Membrane was blocked with 5% milk then incubated with anti-GFP antibody and immunoblots developed with HRP conjugated secondary (sheep anti-rabbit) antibody. Comparable loading was checked by stripping and reprobing the blots with anti-GAPDH antibodies (Santa Cruz Biotechnology, Heidelberg, Germany).

### *GPR156*: Whole mount immunostaining of GPR156 in mouse inner ears

All the animal work was approved by the University of Maryland, Baltimore Institutional Animal Care and use Committee (IACUC 420002). Inner ears were dissected from P10 mice and fixed in 4% paraformaldehyde (PFA) in phosphate buffered saline (PBS) overnight. For whole mount immunostaining, the cochleae were micro-dissected and were subjected to blocking for 1 hour with 10% normal goat serum in PBS containing 0.25% tritonX100, followed by overnight incubation at 4°C with anti-GPR156 antibodies (1:200; Cat#PA5-23857; Thermo Fisher) in 3% normal goat serum with PBS. F-Actin was decorated using Phalloidin (1:300). Confocal images were acquired from Zeiss LSM710 confocal microscope and images were processed using ImageJ software.

## Data Availability

Genetic and phenotypic data for the 100KGP study participants are available through the Genomics England research environment via application at https://www.genomicsengland.co.uk/join-a-gecip-domain. PanelApp gene panels and evidence of associations were obtained using the PanelApp application programming interface (https://panelapp.genomicsengland.co.uk/api/docs/) on the 20th October 2021.

## Code Availability

The rsvr tool and Rareservoir schema is available from https://github.com/turrogroup/rsvr. The BeviMed software package is available from https://cran.r-project.org/web/packages/BeviMed/.

## Acknowledgements

This research was made possible through access to the data and findings generated by the 100,000 Genomes Project. The 100,000 Genomes Project is managed by Genomics England Limited (a wholly owned company of the Department of Health and Social Care). The 100,000 Genomes Project is funded by the National Institute for Health Research and NHS England. The Wellcome Trust, Cancer Research UK and the Medical Research Council have also funded research infrastructure. The 100,000 Genomes Project uses data provided by patients and collected by the National Health Service as part of their care and support. GS was performed by Illumina at Illumina Laboratory Services and was overseen by Genomics England. We thank all NHS clinicians who have contributed clinical phenotype data to the 100,000 Genomes rare diseases programme, and all staff at Genomics England who have contributed to the sequencing, maintenance of the research environment and assembly of the standard bioinformatic files that were required for our analyses. We thank the participants of the rare diseases program who made this research possible. We are grateful to Vaughan Keeley for providing access to paternal DNA (*ERG*), Frances Elmslie for inviting a patient to the clinic (*ERG*), and Thomas Jaworek for technical assistance (*GPR156*). D. Greene was supported by the Cambridge BHF Centre of Research Excellence [RE/18/1/34212] and is supported by Wellcome Collaborative Award 219506/Z/19/Z. V. Hartill is supported by MRC/NIHR Clinical Academic Research Partnership MR/V037617/1. K. Frudd was funded by PG/17/33/32990. D. Pirri is funded by PG/20/16/35047. E. Sackey is supported by Swiss Federal National Fund for Scientific Research n°CRSII5_177191/1. K. Freson is supported by KU Leuven BOF grant C14/19/096. Work at the University of Maryland Baltimore was supported by the NIDCD/NIH grant R01DC016295 to Z. Ahmed.

## Author contributions

D. Greene developed software, conducted analyses and co-wrote the paper. G.E.R.C. provided genetic and phenotypic data and access to the Genomics England research environment. C. Thys performed experiments and interpreted results. B. D. Gelb provided biological interpretation and feedback on the manuscript. K. Freson designed and supervised experiments, provided biological interpretation and contributed to writing the paper. A. Mumford provided clinical oversight, provided biological interpretation and contributed to writing the paper. E. Turro oversaw the study and co-wrote the paper. The following contributions relate to the three gene-specific vignettes. *ERG*: D. Pirri, K. Frudd and E. Sackey performed experiments and interpreted results. S. Mansour and C. L. S. Turner provided additional clinical information. P. Ostergaard coordinated validation and contributed to writing the paper. G. Birdsey designed and supervised experiments and contributed to writing the paper. *PMEPA1*: I. Yamanaka and N. Boeckx conducted experiments and interpreted results. P. Brennan, V. Hartill, J. Harvengt, T. Kosho, M. Masuno and T. Ohata provided clinical information. T. Morisaki and B. Loeys oversaw clinical and experimental studies. H. Morisaki recruited the Japanese cases, conducted experiments, interpreted and analysed results, and oversaw genetic studies. *GPR156*: H. Stewart provided additional clinical information for the compound heterozygous family. K. Taibah clinically evaluated and recruited the p.S642Afs*162 family. A. Giese and K. Ramzan conducted experiments and interpreted results. M. Al-Owain assisted with experiments, interpreted results and contributed clinical information. S. Riazuddin, F. Imtiaz and Z. M. Ahmed designed and supervised experiments, analysed results, and provided reagents and tools.

## Additional information

**Supplementary information** is available for this paper online.

**Correspondence and requests for materials** should be addressed to E. Turro.

## EXTENDED DATA FIGURE LEGENDS

**Extended Data Fig. 1.**
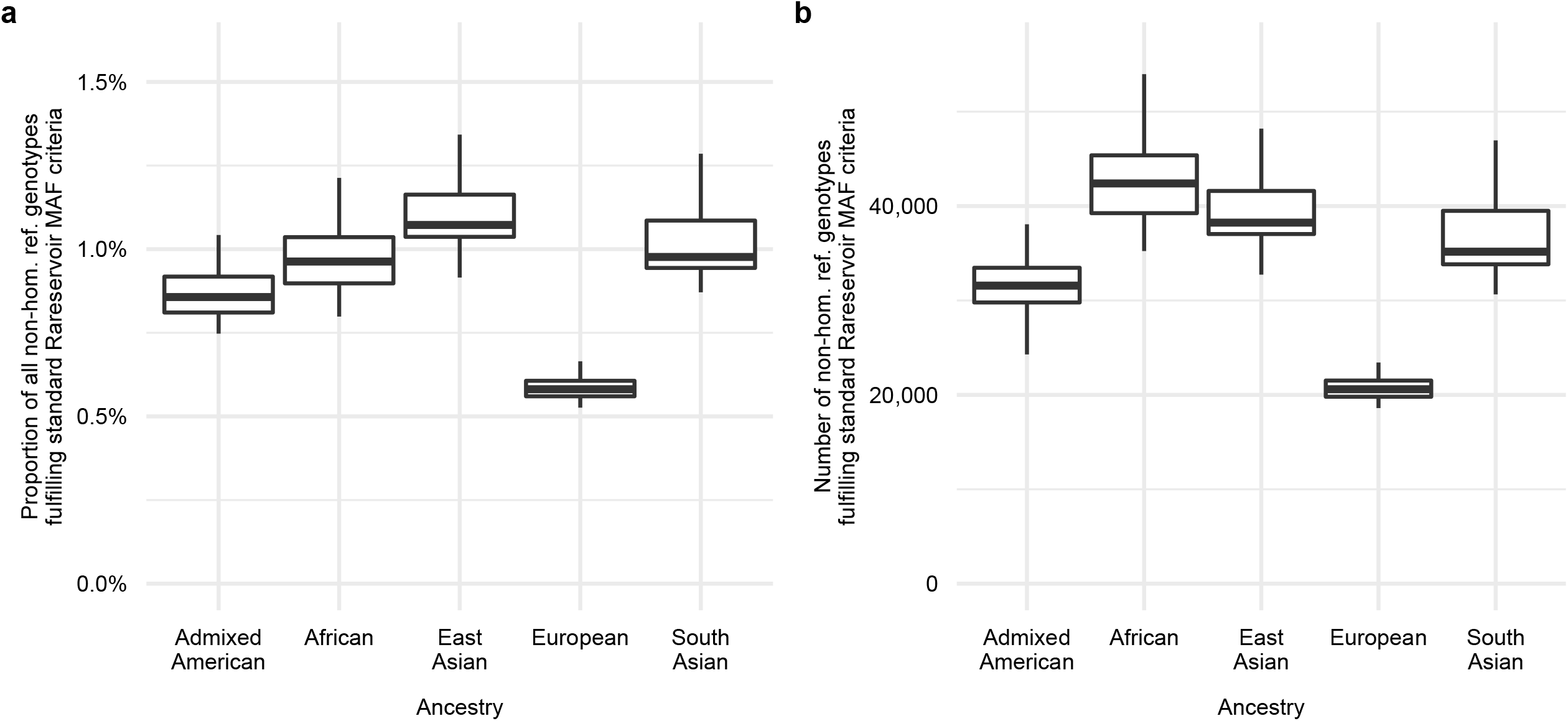
Reduction in the number of genotypes stored per sample. For each of five sets of 100 randomly chosen 100KGP participants with a probability >0.9 of belonging to a specific ancestry group^47^: **a**, boxplots showing the distribution of the number of non-homozygous reference PASSing genotypes for variants on chromosomes 1–22 and X which meet the default Rareservoir MAF filtering criteria (i.e. a PMAF score >0 using gnomAD v3.0 and internal MAF <0.002); **b**, boxplots showing the distribution of the proportion of all PASSing non-homozygous reference genotypes that meet the default Rareservoir MAF filtering criteria. In both plots, the lower, centre and upper lines respectively indicate the lower quartile, median and upper quartile. Whiskers are drawn up to the most extreme points that are less than 1.5× the interquartile range away from the nearest quartile.

**Extended Data Fig. 2.**
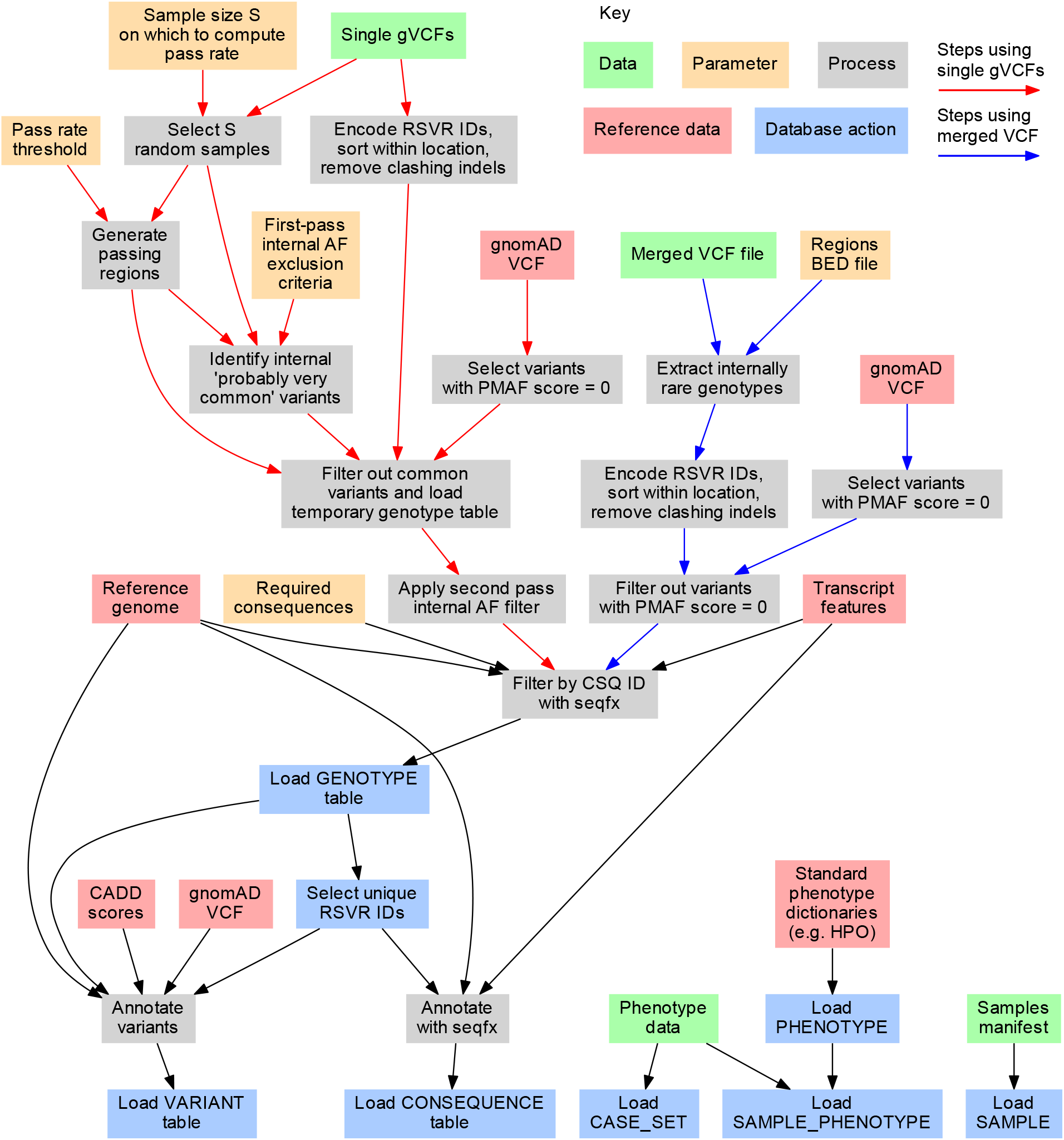
Detailed schematic of the database build procedure. Variants may be imported to a Rareservoir from either single gVCF files or a merged VCF file, following the procedures indicated by red and blue arrows respectively.

**Extended Data Fig. 3.**
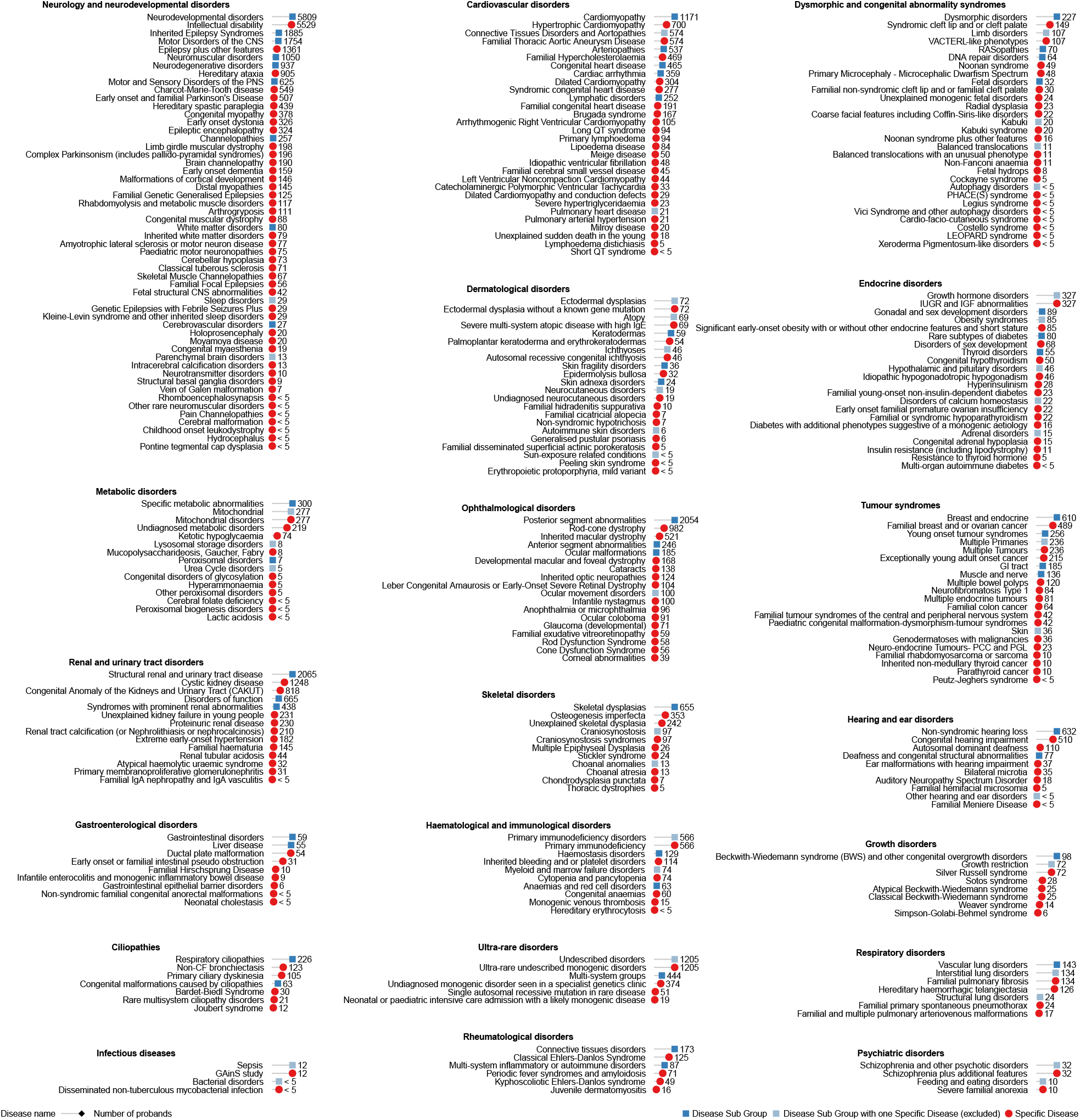
The 269 case sets. The names and sizes of the case sets used for the genetic association analyses, grouped by Disease Group and coloured by type (Disease Sub Group or Specific Disease). Disease Sub Groups with only one Specific Disease were excluded to avoid repeating identical analyses. Case sets smaller than 5 are labelled ‘<5’ and shown as having size 4 to comply with 100KGP policy on limiting participant identifiability.

**Extended Data Fig. 4.**
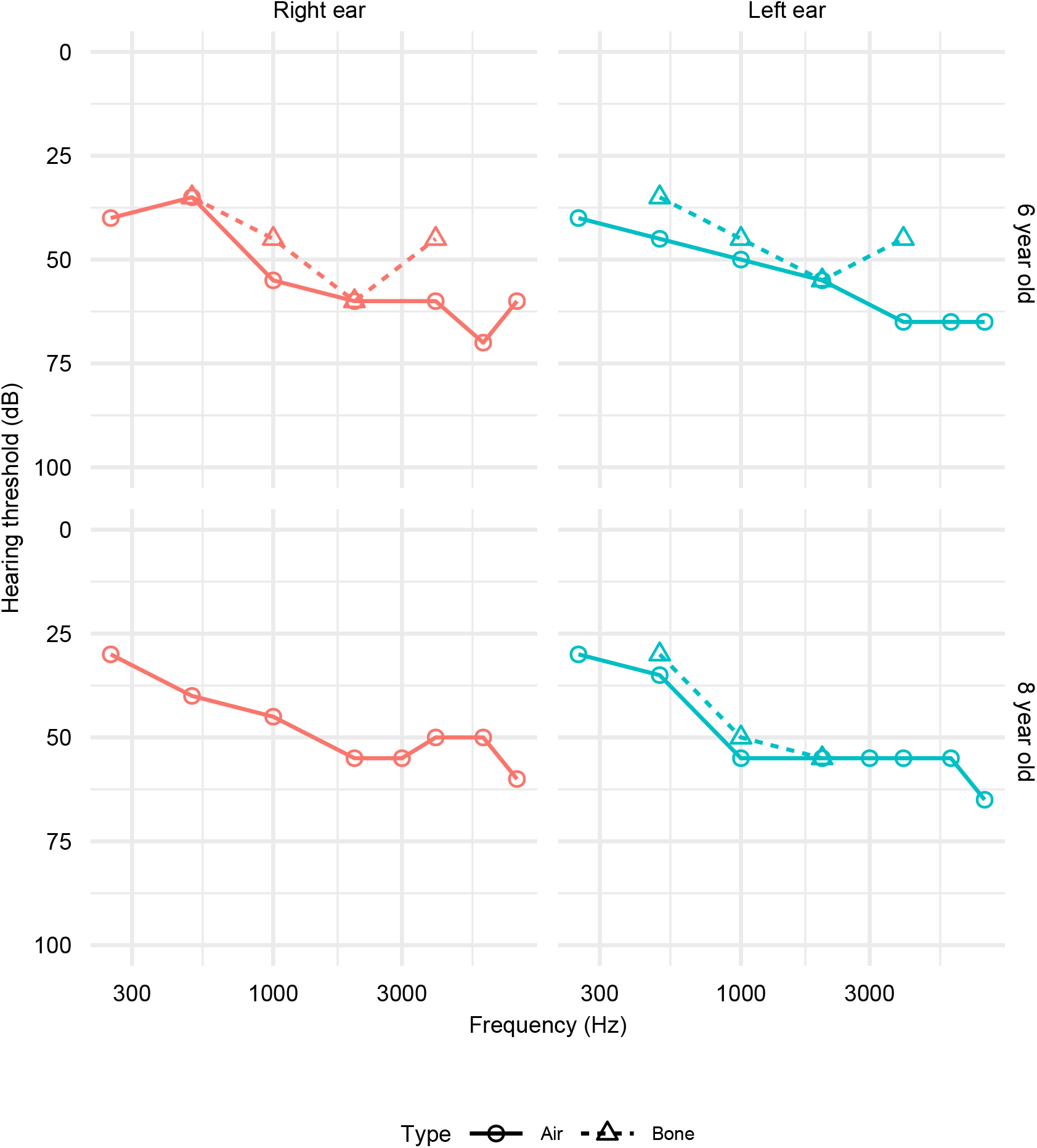
Illustrative audiograms for *GPR156* cases. Air and bone conduction audiograms for the two affected daughters of the family with compound heterozygous *GPR156* truncating alleles.

## SUPPLEMENTARY TABLE LEGENDS

**Supplementary Table 1.**
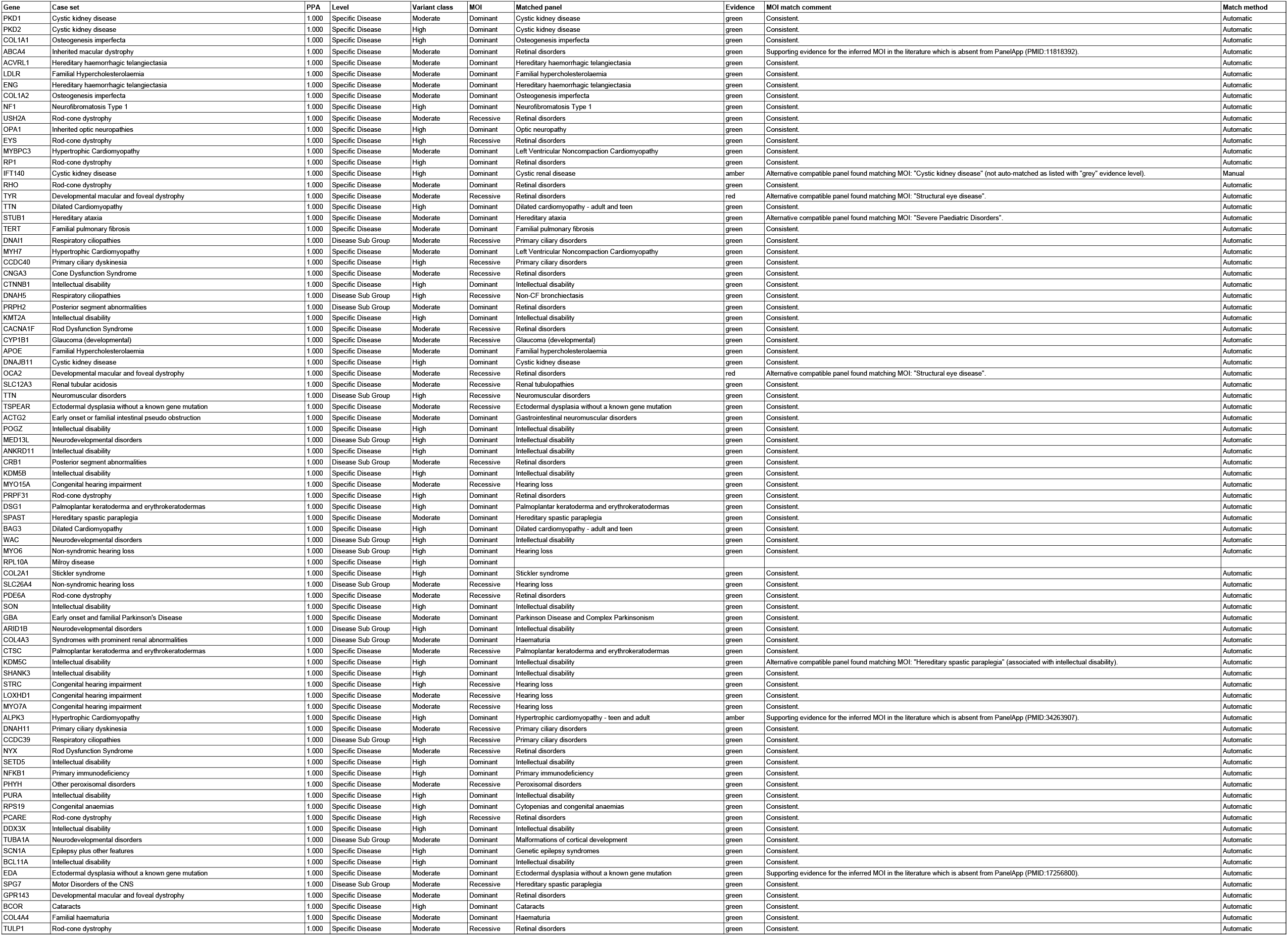
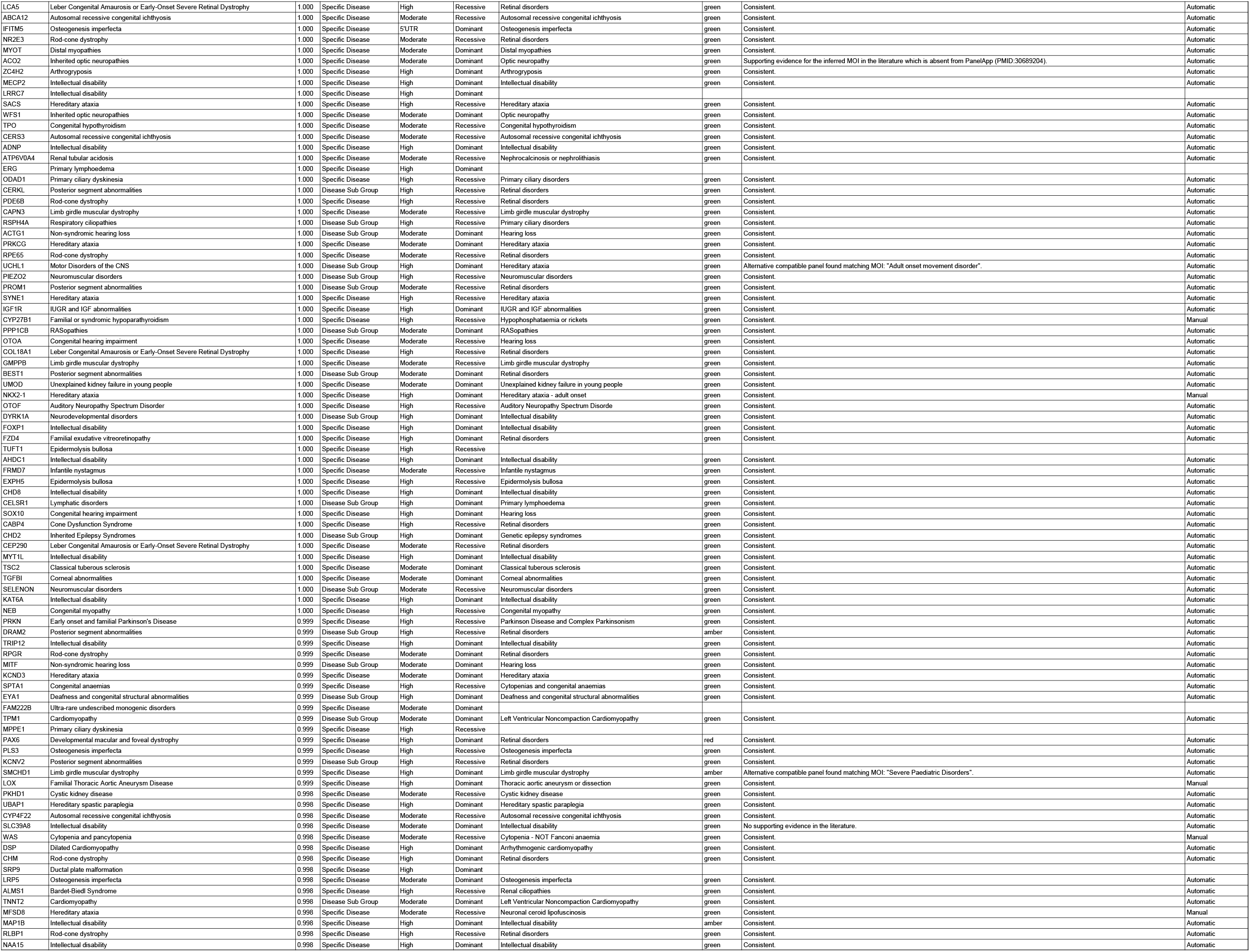
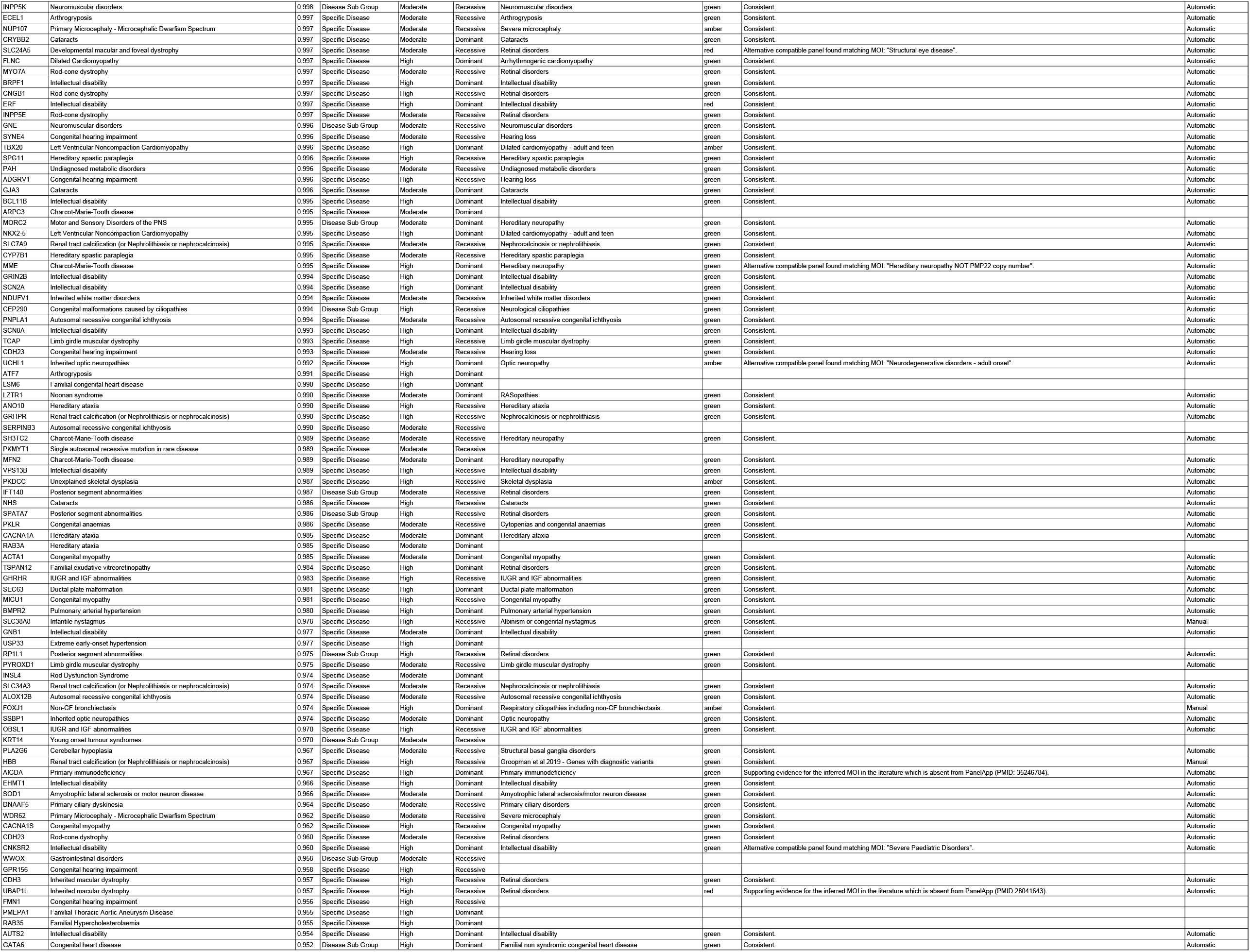
Detailed annotated results of the genetic association analyses. Table of associations shown in Fig. 2 annotated with BeviMed PPAs (PPA), the level of the case set in the disease label hierarchy (Level), the inferred variant class and MOI for the association, the matched PanelApp panel for the association and associated evidence level, notes on the consistency between the MOI listed by PanelApp for the association and the inferred MOI (MOI match comment), and the method that was used to find the match (Match method, either ‘Automatic’ or ‘Manual’).

